# Leveraging community-centric digital health technology to sustain healthier living – inferences from a wearables-based exergaming program

**DOI:** 10.1101/2025.08.12.25332025

**Authors:** Sai G.S. Pai, Kai Zhe Tan, Huanyu Bao, Ben Pham Phat, Yin-Leng Theng, Edmund W. J. Lee, Navrag B. Singh

**Affiliations:** Singapore-ETH Centre, Future Health Technologies Program, CREATE campus, 1 CREATE Way, #06-01 CREATE Tower, Singapore 138602; Laboratory for Movement Biomechanics, Institute for Biomechanics, Department of Health Science & Technology, ETH Zürich, Zürich, Switzerland; Wee Kim Wee School of Communication and Information, Nanyang Technological University, Singapore; Department of Media and Communication, City University of Hong Kong, Hong Kong SAR

**Keywords:** Digital health technology, exergames, mobile health applications, wearables, active ageing, older adults, adherence, physical activity, well-being

## Abstract

In this study, the development of a community-centric exergame-based program to improve the well-being of older adults and promote adherence to a healthy lifestyle is explored. Thirty participants competitively engaged in three exergames targeting upper and lower limb movements, over four weeks, at a community center. An app, SingaporeWALK, was developed to track and visualize movement data from wearables and self-reported well-being scores. All participants reported increased motivation to use digital health technologies (DHTs), along with improvements in physical, emotional, and social well-being. Wearable data revealed week-to-week differences in movement characteristics, especially between weeks 1 and 2 suggesting a learning effect. Participants who performed better at exergames exhibited more varied movements, indicating ability to learn, alter, and optimize movement over the weeks. The study demonstrates that community-based exergaming is effective in enhancing overall well-being, while increasing interest in active ageing by leveraging DHTs and has potential to support targeted interventions.

## 1. Introduction

By 2030 1 in 6 people will be aged 60 years or above, while the global older adult population is expected to reach 2 billion in 2050 (Johnson, 2008a; Organization, 2022). Increased life expectancy has contributed to an ageing society (Johnson, 2008b), however, the proportion of life spent in good health has stagnated (from 15.1 in 2010 to 15.3 in 2020 globally,World Health Organization, 2024b). The implications of longer life in poor health are profound (Calder et al., 2018). Healthy ageing (World Health Organization, 2024a), the sustenance of physical, social, and emotional well-being, can alleviate this challenge, thereby reducing incidences of non-communicable diseases and economic burden on the health care system (Crombie, 2004; Calder et al., 2018). While diverse physical activity-based programs have been developed, an equitable holistic, unified, and scalable approach that targets the multi-factorial nature of healthy aging is still missing (McPhee et al., 2016; Bao et al., 2024a). In addition, motivation toward acceptance and continued adherence has been a challenge (Larsen et al., 2013; Koh et al., 2022; Guu et al., 2023).

Exercises in the form of games – exergames –motivate older adults to be physically active and to improve physical function as well as cognitive abilities (Kappen et al., 2019). Exergaming in social and community-based settings, *social exergaming* (Lin et al., 2023; Chan et al., 2024), such as playing the games either collaboratively or competitively under multiplayer settings, has been shown to increase motivation(Li et al., 2021), improve social and emotional well-being (Zheng et al., 2020a), and reduce loneliness (Strand et al., 2014). Social exergaming interventions can enhance adherence to behaviours crucial for healthy ageing (Li et al., 2021).

Digital health technologies (DHTs) such as mobile applications and wearables, can enable older adults to easily and continuously measure, monitor, and relay pertinent health metrics in real time for personalized interventions and mitigate challenges in promoting healthy ageing (Chen et al., 2023). However, these technologies must be carefully designed (Guu et al., 2023) and integrated with intervention programs to inform progress and goals to achieve and evaluated to benefit public health (Money et al., 2024), while accounting for barriers to use such as lack of affordability and accessibility, deficit in trust and technology anxiety (Koh et al., 2022; Frishammar et al., 2023). In this manner, a socially viable gaming-based ecosystem can facilitate the acceptability and adherence of DHTs and physical activity-based programs for healthy ageing.

This study implements a community-centric DHT, comprising wearables and a mobile application with a dashboard, to enhance motivation and adherence to an exergaming program for older adults. The exergaming program adopted a participatory approach where participants competed against each other over four weeks (training period) while playing exergames (Artic Punch, Fruit Ninja and Piano Step) with their movements recorded via the use of wearables. The program culminated in a two-week competition phase: during the 5^th^ week, participants engaged in semi-finals to shortlist finalists, followed by a final Fun-Day event in 6^th^ week. This competitive structure served as motivation for participants to improve their gameplay performance throughout the program.

In this study, we aim to evaluate the acceptance and motivation of older adults towards such a community-centric, DHT-enhanced solution for supporting healthy ageing. This is facilitated by self-reported information from participants before and after the completion of the four-week training period. We also aim to evaluate exergaming performance during the program to assess its potential for use in targeted interventions. The performance assessment was carried out using the movement data collected with wearable inertial measurement unit (IMU) sensors during the training period.

## 2. Methods

### 2.1. Study design and participants

This study is a community-based investigation on healthy ageing benefits of participatory exergaming programs. Individuals were assessed longitudinally, once a week for 6 weeks, while playing interactive games at a local community center. The study culminated in a ‘Fun-Day’ competition event during the 6^th^ week, with participants selected through a mini-competition held during the 5^th^ week, following a four-week training period. Movement and health data was collected from older adults participating in exergaming activity over the four-week training period (see Figure 1-a). Ethical approval for this study was granted by the Nanyang Technological University – Institutional Review Board (IRB-2022-734). Written informed consent was obtained from all participants prior to their involvement in the study. All procedures adhered to the Declaration of Helsinki.

**Figure 1.**
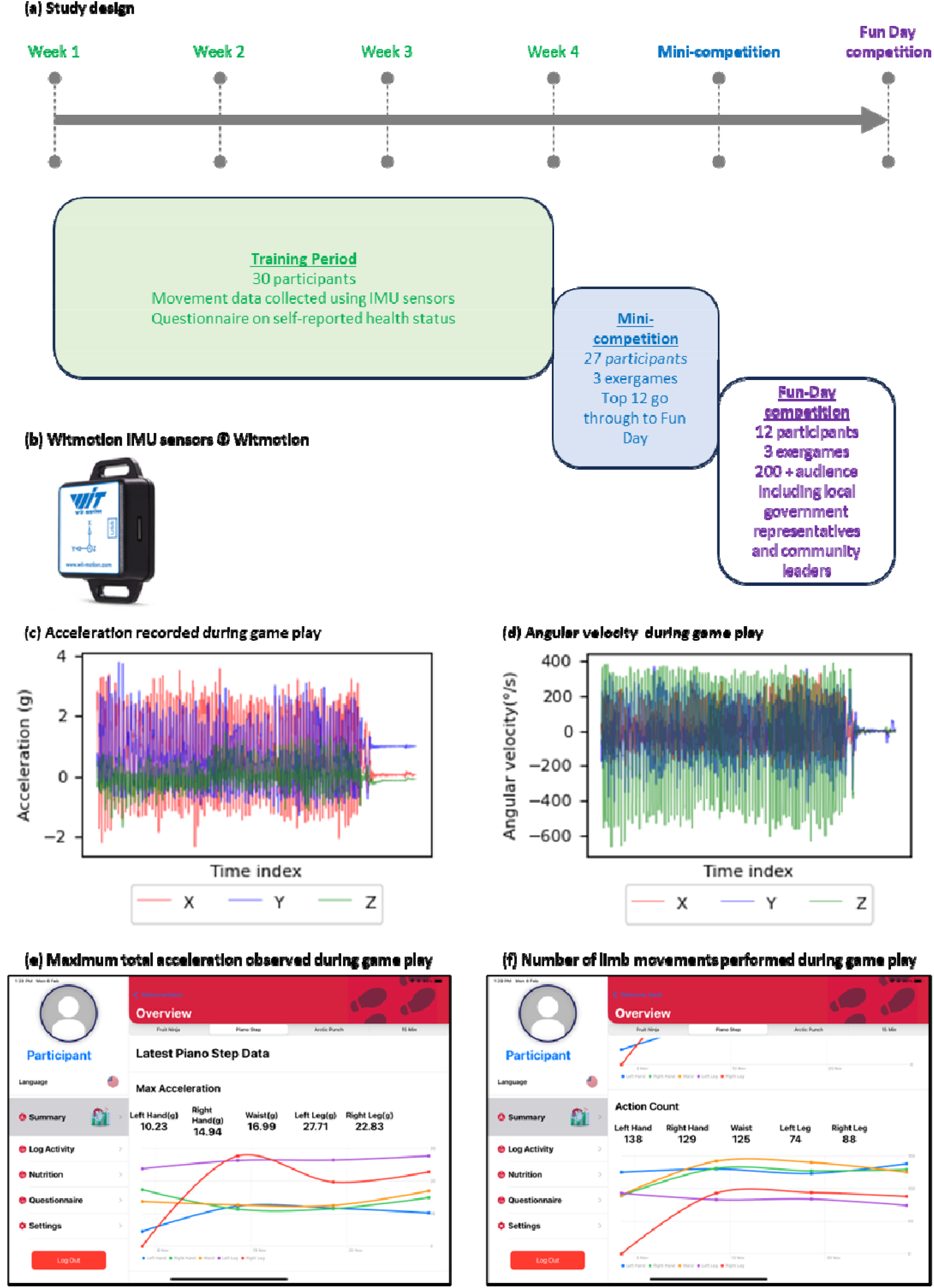
(a) Schematic of the data collection study conducted at a community centre. The study involved a data-collection period (training period) of four weeks followed by a mini-competition and a “Fun-Day” Competition to enhance the socialization and encourage adherence to healthy ageing. (b) Witmotion IMU sensor. (c) & (d) Triaxial acceleration and angular velocity recorded with an IMU sensor during game play. (e) and (f) Summary sensor features that are processed immediately after game play using algorithms embedded within a custom-designed app and shared with participants as feedback.

During the four-week training period, 30 older adults (25 females; mean age 73.23 years with a standard deviation of 7.33 years) participated in exergames every week. Participants were asked to play three exergames, Arctic Punch, Fruit Ninja, and Piano Step. These games were chosen based on their similarity with exercises recommended for promoting an active lifestyle among older adults in Singapore (Synapxe Pte. Ltd, 2024). The sample size (n = 30) for this study was chosen based on the feasibility and practicality of running an interactive exergaming program with the community centre that supported the six-week program.

Participants who completed the four-week training period took part in a mini-competition (Week 5), where they played the same three exergames from the training period. The top 12 participants assessed based on game-play scores qualified for the final Fun-Day competition and are categorized in this study as Group A. The remaining 15 participants who did not qualify for the competition are denoted in this study as Group B. The Fun-Day was a celebratory event organized by the community centre that put together tech-centric and traditional games. Older adults competed on the day, playing the three exergames, with the audience comprising of the local community, friends and family providing verbal encouragement.

### 2.2. Study Procedures and data collection

Each game session lasted 2 to 3 minutes and was played twice. The initial gameplay served as a trial session for participants to familiarize themselves with the game’s rules. The subsequent round included recording movement patterns during gameplay using wearables integrated within SingaporeWALK (**W**earables-based **A**pp for community **L**iving to enhance **K**nowledge and awareness of mobility) mobile application.

During the four-week training period, 20 participants were quasi-randomly assigned to play these games with a peer (10 pairs) and the remaining 10 participants played with a health coach. A health coach is a volunteer on-site who works with the participants to review their scores, and to work collaboratively with participants to identify areas of improvement. During these four weeks, participants, played three exergames while being monitored with wearables and answered questionnaires on emotional, social, and physical well-being as well as outlook and motivation towards physical activity and the role of technology. During the mini-competition and Fun-Day, participants did not use wearables and the SingaporeWALK app.

Wearable inertial measurement units (IMUs - WT901BLECL-MPU9250 Witmotion Shenzhen Company Limited, China; see Figure 1-b) are used in this study to record movement of participants. These miniature-sized wearables are embedded with a triaxial accelerometer (range ±16g), a gyroscope (±2000degrees/s), and a magnetometer. The data was collected at a sampling rate of 50Hz. During the four-week training period, participants were fitted with five wearable sensors during gameplay. The sensors were strapped to the participants at their wrists (left and right), waist, and ankle (left and right) and allowed data capture (acceleration as shown in Figure 1-c and angular velocity as shown in Figure 1-d) in an unobtrusive manner.

SingaporeWALK is an application that has been developed to promote health and well-being, specifically targeting older adults. It serves as a data collection platform and a visualization and communication tool for metrics that promote healthy ageing. Participants and health coaches can utilise this visual feedback to make healthier lifestyle choices. The app comprises of physical activity, nutrition and well-being trackers. This app also enables data recorded by the wearables to be relayed to an iPad via Bluetooth^®^ and stored locally.

SingaporeWALK app enables analyzing wearables-data in near real-time basis to provide visual feedback (Figure 1-e and Figure 1-f) to participants/health coaches related to mobility outcomes such as the number of limb movements performed during the game and speed of movement. Participants are encouraged to discuss these metrics and their performance with peers, which serves as motivation to improve at gameplay, while simultaneously creating avenues to socialize, as well as adhere/sustain to and even improve upon physical activity goals.

### 2.3. Data processing algorithms

Acceleration data from the wearable sensors is acquired as a time-series (see Figure 1-c). Using triaxial acceleration data, the total acceleration is calculated using Eq. (1).

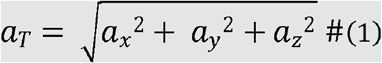

In Eq. (1), *a*_*x*_, *a*_*y*_ and *a*_*z*_ are the acceleration time series along x, y and z-axis respectively of the wearable data and *a*_*T*_ is the total or resultant acceleration. Using Eq. (1), the total acceleration is calculated independently for data collected from each wearable (wrists, waist, and ankle). The maximum value of the total acceleration (see Figure 1-e) and the number of cycles observed in the total acceleration time series (names as “Action Count”; see Figure 1-f) at each sensor location was provided as feedback to the participants with the SingaporeWALK app after each gameplay.

Angular velocity data obtained using a gyroscope (see Figure 1-d) is analysed to gain further insights into movement characteristics captured during gameplay. Specifically, range-of-motion (ROM) and peak angular velocity (AV) are calculated for each movement cycle during gameplay. ROM and AV may be informative of a participant’s mobility, repertoire (variability in movement) and speed. Information regarding ROM and AV was not integrated into the SingaporeWALK for live feedback. This post-processing of angular velocity data has been performed with Python3 (Python Software Foundation, USA, https://www.python.org/).

The procedure for analysing angular velocity data from a gyroscope to obtain ROM and AV is shown in Figure 2 and has been adapted from Warmerdam and coworkers (Warmerdam et al., 2020). The primary modification includes accounting for differences in movement signatures between walking and playing exergames. The tri-axial raw angular velocity data from the sensor is low-pass filtered (fourth-order, zero-phase lag, Butterworth, 10 Hz cut-off frequency) to remove high-frequency noise and other movement artefacts. Principal component analysis is then performed on the filtered, tri-axial limb angular velocity data during gameplay to identify the primary direction of limb movement (first component). This component (time series) is further integrated numerically to obtain angles from angular velocity.

**Figure 2.**
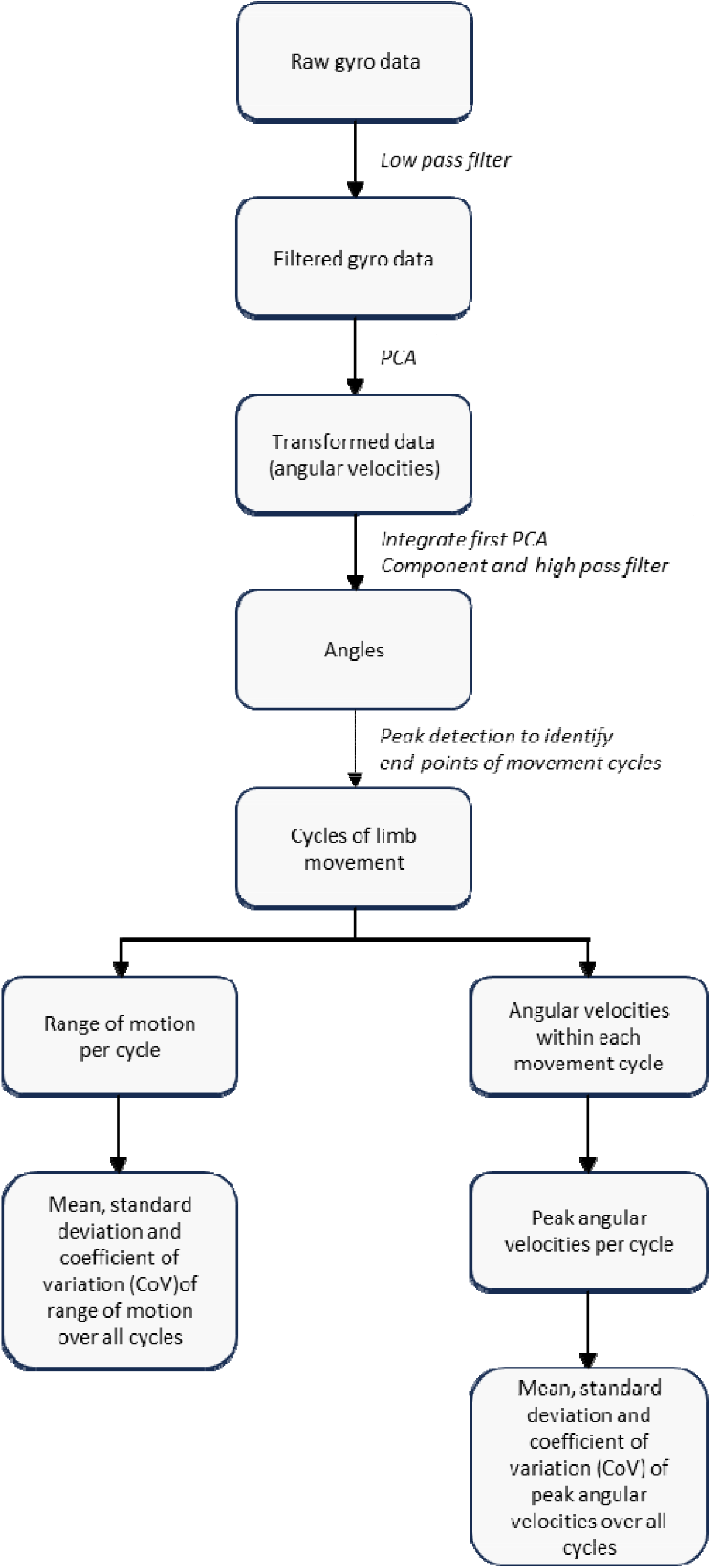
Steps involved in processing wearables data to extract metrics such as mean range of motion and mean peak angular velocity recorded during a session of gameplay.

The integrated angular time series is then high-pass filtered (fourth-order, zero-phase lag, Butterworth, 0.2 Hz cut-off frequency) to remove low-frequency noise introduced by numerical integration (drift etc.) approach. The time series thus derived corresponds to an estimation of the limb movement angles during gameplay.

### 2.4. Outcomes

The primary outcome was the difference between perception of health technology use and interest to exercise between Week 1 and Week 4 of the training period. This information was assessed based on a battery of questionnaires completed by participants at Week 1 (pre-test) and Week 4 (post-test). The questionnaires include (a) perceptions of health technology use (wearables, application, exergames), (b) exercise intentions (motivation, interest, etc.) and (c) physical and mental health status. Additionally, participants completed a short weekly survey on (a) self-reported physical health, (b) psychological, (c) social, and (d) emotional well-being.

Additionally, movement of participants during gameplay were recorded using wearables. Participants move their limbs to achieve a certain objective within each game, such as slicing the fruits on screen while playing “Fruit Ninja”. Each swing corresponds to a pair of extrema (maxima and minima) in the time series of angles calculated. Within each swing (cycle of movement), the corresponding characteristics of ROM (degrees, °) and AV (degrees per second (°/s) are evaluated.

Within a cycle of movement, the ROM of a swing is evaluated as the difference between the starting and ending angle (difference between the extrema). During a gameplay session, if the participant swings ‘*N*’ times, then the mean of ROM is assessed over the *N* swings. This mean of ROM is evaluated for each sensor location. Similarly, the mean of AV is assessed as the average of the peaks of angular velocities observed over *N* cycles recorded within one session of gameplay for each participant.

The coefficient of variation (CoV) for movement characteristics, ROM and AV, is evaluated as the ratio between the standard deviation and the mean of the movement characteristic for one session of gameplay. Movement characteristics from specific sensors assessed for each game are reported in Table 1.

**Table 1.** Details regarding sensor location from which movement data has been evaluated for each game.

| **Game** | **Characteristics calculated** | **Limbs** |
| --- | --- | --- |
| Arctic Punch | Mean and CoV of ROM and AV | Arm swing movements (right and left wrist sensors) |
| Fruit Ninja | Mean and CoV of ROM and AV | Arm swing movements (right and left wrist sensors) |
| Piano Step | Mean and CoV of ROM and AV | Foot movements (right and left ankle sensors) |

### 2.5. Statistical analysis

Significance in differences between movement characteristics over the weeks as well as pre-and-post study responses to questionnaires were assessed with the Wilcoxon Signed-Rank Test, with threshold at *p*<0.05 for statistical significance. The significance tests are reported in Section D and Section E in the Supplementary Materials.

## 3. Results

Over the four-week training period, a general increase in participant’s interest was observed in exercising as well as the use of DHTs for this purpose. In Table 2, the change in participants’ self-reported interest in exercising and using digital health technologies is reported (with standard error reported in the brackets). This increase in interest towards the use of technology for exercising was consistent across the two populations, the ones who qualified for the final week Fun-Day competition (Group A) as well as those who didn’t qualify (Group B). The participants were overall significantly (*p*<0.05) interested in exercising, using wearables and digital health apps with improved perception of these apps compared with before the study. The *p-value* assessment is carried out comparing the absolute values of response pre- and post-study. The absolute values along with p-values are reported in Supplementary Materials, Section E.

**Table 2.** Mean of change in reported interest in exercising and use of digital health technologies by participants before and after the study for the two groups (those who qualified for the Fun-Day event, Group A, and those who did not, Group B). * Denotes significant difference (*p*<0.05) in pre and post study responses. Standard error reported in “(*brackets*)”.

| For responses related to | Percentage (%) change for |  |  |  |
| --- | --- | --- | --- | --- |
|  | Group A (n=12) | Group B (n=14) | Overall (n=27) |  |
| Interest in exercising | 32.25 (10.56) * | 31.31 (8.64) * | 31.75 | (6.50) * |
| Interest in exergaming | 7.53 (5.75) | 9.33 (7.64) | 8.50 | (4.64) |
| Interest in wearables | 17.48 (10.55) | 4.46 (4.20) | 10.47 | (5.45) * |
| Interest in digital health apps | 38.43 (8.47) * | 32.88 (7.27) * | 35.44 | (5.36) * |
| Perceived utility of health app | 21.48 (11.27) | 32.40 (17.18) * | 27.36 | (10.08) * |

Additionally, over the four weeks of training, only 3 out of 30 participants (dropout 10%) dropped out. Reasons for dropouts were personal commitments and scheduling conflicts rather than any averseness to the study.

Furthermore, data from the four-week training period, analysed as described in Section 2, showed considerable differences. These differences are shown in Figure 3. The movement characteristics of participants changed drastically from Week 1 to Week 2. The distribution of the movement characteristics (ROM and AV) for Week 2 has lower variability compared with Week 1 for all three games (Figure 3 (i) – a, b, and c). The mean and variability of movement characteristics for Week 2 is up to 60% less than in Week 1. Differences mean of ROM and AV between Week 1 and Week 2 is significant (*p* < 0.05). These differences are likely due to participants familiarising themselves with the game as well as associated technology (wearables, SingaporeWALK app etc.). No such differences were observed in subsequent weeks, likely due to an improved understanding of gameplay.

**Figure 3.**
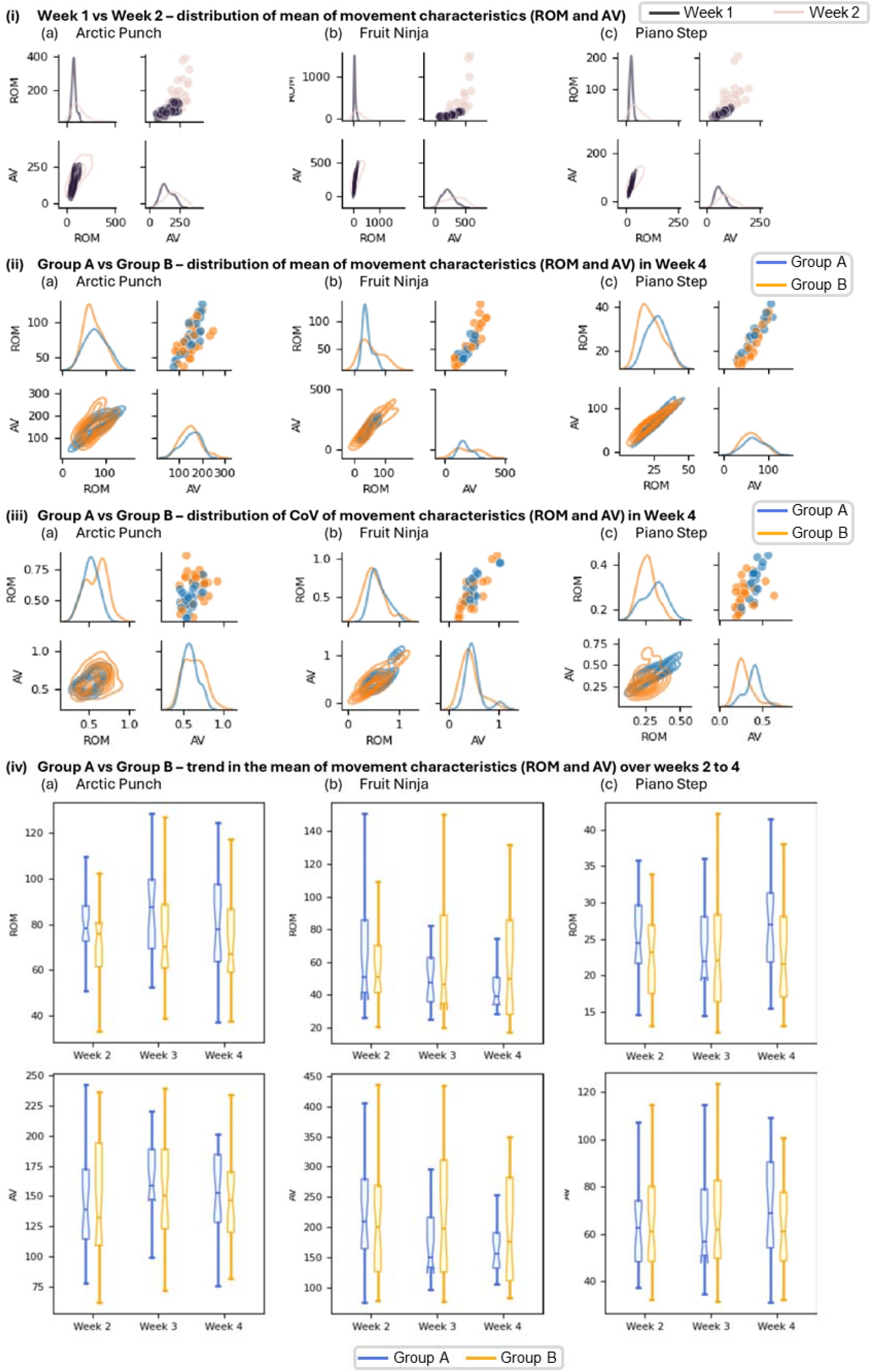
(i) Change in movements of participants from week 1 to week 2 as they learn to play the three games (a, b and c), (ii) Distributions of mean of movement characteristics of participants from Group A and Group B, (iii) Distributions of CoV of movement characteristics of participants from Group A and Group B and (iv) Week-by-week of evolution of mean of movement characteristics of participants from Group A and Group B. Units for ROM is degrees (°) and AV is °/s

**Figure 4.**
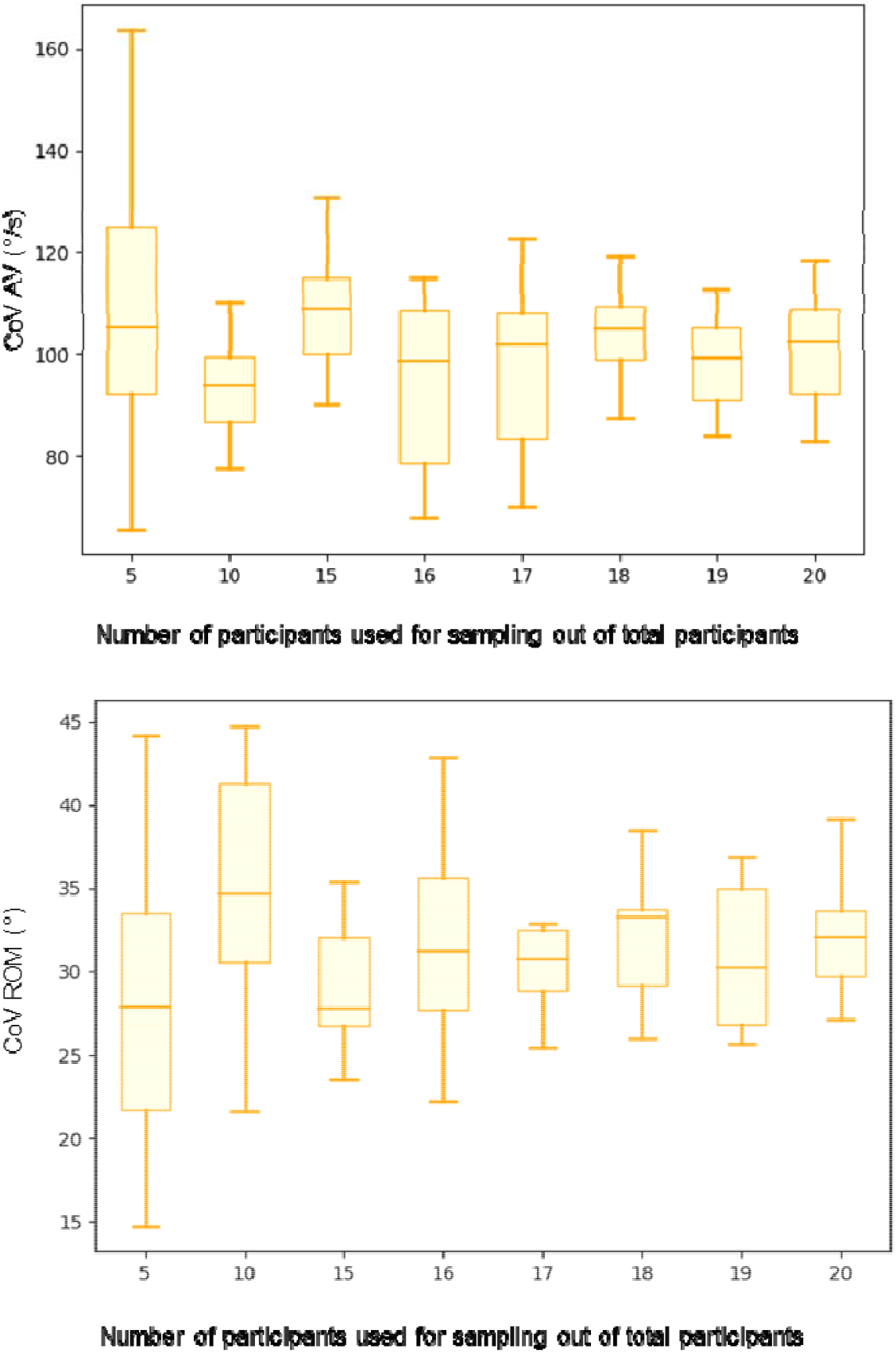
Mean value of the distribution for CoV of AV and ROM of participants while playing Fruit Ninja begins stabilizes when calculated with more than 15 participants.

From Week 2 onwards, no overall difference in movement characteristics was observed. However, variations in movement characteristics were observed between individuals. This difference was prominent between two groups: Group A - those who participated in the mini-competition and qualified for the Fun-Day and Group B - those who did not qualify for the Fun-Day event. In Figure 3 (ii) and Figure 3 (iii), the differences in movement characteristics, ROM and AV, respectively, from Week 4 of participants in groups A and B are shown. Observations from Figure 3 (ii) and Figure 3 (iii) are reported in Table 3.

**Table 3.** Observations on movement characteristics and their differences in the pool of participants within Groups A vs B in the final week of gameplay (Week 4) at the end of exergaming training. *p-values* for significance in differences are reported in Supplementary Materials, Section D.

| Game | Observation |
| --- | --- |
| **Arctic Punch** | Group A participants exhibited a greater mean of ROM and AV compared to Group B (up to 8% greater; see Figure 3-ii-a).<br>The within or intra-participant variations, CoV, in ROM and AV, for Group A was lower than for Group B (up to 13% lower; see Figure 3-iii-a). |
| **Fruit Ninja** | No drastic difference was observed in the mean of the movement metrics (see Figure 3-ii-b). However, variances of mean movement values of Group A participants (inter-participant variance) were considerably lower than those from Group B (variance of the plot in Figure 3-ii-b; up to 75% lower).<br>Greater within-participant variability was observed in the movement (ROM and AV) of Group A participants compared to Group B participants (up to 15% greater; see Figure 3-iii-b). ROM variability between Group A and Group B participants is significantly different in Week 4. |
| **Piano Step** | The Group A participants demonstrated greater speed and range of movement to press down on highlighted keys, even if they were far away from the current standing location (up to 15% greater; see Figure 3-ii-c) compared with participants from Group B.<br>Group A participants showed significantly greater movement variability (up to 25% greater; see Figure 3-iii-c), i.e., they were likely to press down on keys far away from them compared with Group B participants. |

In Figure 3 (ii) and Figure 3 (iii), movement characteristics from gameplay during Week 4 were compared to assess differences between the movement of participants within Groups A and B. ROM variability differences between groups A and B are significant (*p*<0.05) in Week 4. In Figure 3 (iv) – (a), (b), and (c), the progression and changes in movement characteristics of both groups of participants over the last three weeks for the games, Arctic Punch, Fruit Ninja, and Piano Step, respectively, are shown. The median of movement characteristics for Group B participants did not change drastically over the weeks. This observation is consistent across the three games. Conversely, for Group A participants, the median of the movement characteristics varied drastically week-by-week (up to 50% change from Week 2 to Week 4; see Figure 3-iv-b).

Data on participants’ health (physical, emotional, social, and psychological well-being) were collected from participants before and after the four-week training period using a questionnaire. In Table 4, the change in participants’ self-reported health is presented (with standard error reported in the brackets). Both groups of participants, irrespective of their performance in gameplay, reported improvements in their health scores. The *p-value* assessment reported in Table 4 is carried out comparing the absolute values of response pre- and post-study. The absolute values along with *p-values* are reported in Supplementary Materials, Section E.

**Table 4.**
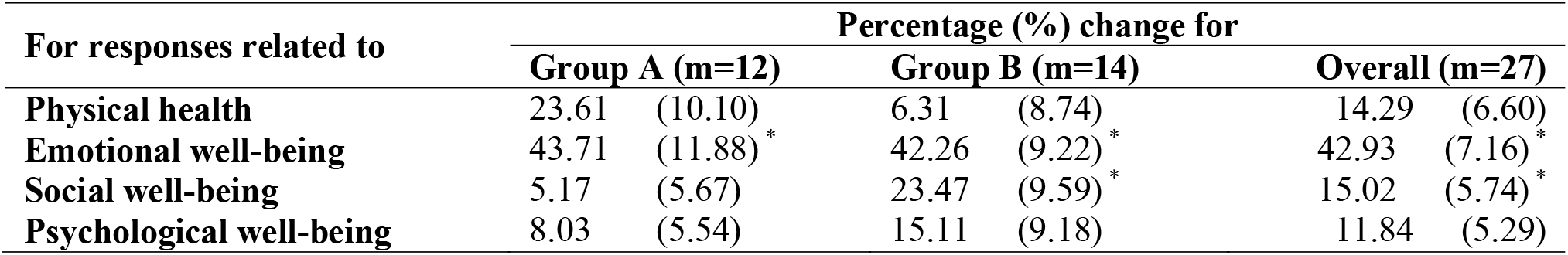
Change in reported health scores by participants before and after the study for the two groups (those who qualified for the Fun-Day event and those who did not). * Denotes significant difference (*p*<0.05) in pre and post study responses. Standard error reported in “(*brackets*)”.

| For responses related to | Percentage (%) change for |  |  |  |  |  |
| --- | --- | --- | --- | --- | --- | --- |
|  | Group A (m=12) |  | Group B (m=14) |  | Overall (m=27) |  |
| Physical health | 23.61 | (10.10) | 6.31 | (8.74) | 14.29 | (6.60) |
| Emotional well-being | 43.71 | (11.88) * | 42.26 | (9.22) * | 42.93 | (7.16) * |
| Social well-being | 5.17 | (5.67) | 23.47 | (9.59) * | 15.02 | (5.74) * |
| Psychological well-being | 8.03 | (5.54) | 15.11 | (9.18) | 11.84 | (5.29) |

## 4. Discussion

This research aims to address gaps in research on developing a holistic and equitable digital technology-based program for engaging and motivating older adults to be physically active(Bao et al., 2024b). For this purpose, an integrated community-centric program, first of its kind, was conducted to educate and motivate the elderly to exercise in a competitive and gamified manner while leveraging DHTs (wearables and SingaporeWALK). Our results demonstrate that the participatory and gamified use of DHTs, along with near real-time feedback on gameplay provided with wearables-based analytics improved the motivation of older adults to be physically active as well as their emotional, social, psychological, and physical health.

Older adults were found to be more interested and motivated to exercise and use digital health technology (SingaporeWALK app and wearables) (see Table 2) after taking part in this pilot program. Near real-time feedback was also provided to participants about their gameplay. This, in addition to the low dropout rate, augments the proposition that older adults demonstrate high adherence, interest, and motivation to physical activity and use of digital health technologies when experienced in a gamified and community-centric participatory environment (Wu et al., 2015; Li et al., 2018). Also, as reported in Table 2, the perceived utility of the health app improved in older adults after using the application in a gamified and community-centric manner. This also suggests that apps and wearables should be designed to encourage social and community engagement.

Performing gamified physical activity improves the physical, social, emotional, and psychological well-being of older adults (Hall et al., 2012; Larsen et al., 2013; Zheng et al., 2020b), especially when performed in a community-centric manner (Brox et al., 2011). Our observations are in line with these studies. The positive outcomes, as reported in Table 4, affirm that the DHTs developed within this study help older adults to be physically active, thereby improving their social, emotional, and psychological well-being as well. Particularly, emotional well-being improved by greater than 35% over the four-week period, which may primarily be due to the community-centric aspect of the exercise (Brox et al., 2011).

The use of wearables enabled assessment of movement in older adults participating in exercises in the form of games (exergames) in an unobtrusive manner. This consequently helped assess the effect of fast learning (difference in movement from week 1 to 2) vs slow learning (from week 2 to week 4) (Karni et al., 1998; Doyon and Benali, 2005) processes, as shown in Figure 3(i). Fast learning involves familiarizing and selecting and establishing an optimal routine for performing a task (Sarwary et al., 2018a). While, slow learning reflects the ongoing long-term, structural, modifications of basic motor modules (Sarwary et al., 2018b). Therefore, moving forward, it is important to account for motor learning effects while using digital technologies to assess mobility-related health outcomes (Shcherbina et al., 2019), such as cardio-vascular risk assessment (del Pozo Cruz et al., 2022) and for rehabilitation after injury or illness.

Wearables-enabled assessment of movement and consequently objective feedback on movement strategies adopted as well as gameplay performance can be a motivating factor to encourage older adults to be physically active. For the Arctic Punch game, participants that moved faster (higher AV, see Figure 3-ii-a), with a greater range of motion (higher ROM, see Figure 3-ii-a), and did so consistently (lower variation, see Figure 3-iii-a) played the game better (Group A versus Group B). However, this observation was not consistent across the games.

For Fruit Ninja, the differences in mean values of movement characteristics among those who played the game better are low (variance of the probability distribution of the mean of ROM and AV shown in Figure 3-ii-b). This indicates adaptation towards a movement strategy that optimises speed and range of motion for effective gameplay. Additionally, contrary to the observation made for the Arctic Punch game, greater variability (intra-participant variability; higher median of CoV distribution) was observed in movement characteristics (ROM and AV) of Group A participants compared to their Group B counterparts (see Figure 3-iii-b). In the Piano Step game, participants that demonstrated greater repertoire (variability, see Figure 3-iii-c), speed and range of movement (see Figure 3-ii-c) to press down on highlighted keys, even if the keys were very far away from the player location, played the game better. In future, choice on the type of exergame could be customized to the needs of the participants or patients. As both Fruit Ninja and Arctic Punch predominantly require upper body motion wheelchair bound participants might be able to partake in the rehabilitation program.

The ability to learn, alter, and optimize movement over the weeks of training was also a key factor in enhanced gameplay (greater variability in weekly median values of movement characteristics of Group A participants compared with Group B as shown in Figure 3-iv). Each game required a specific movement strategy focusing on different combination of limbs vs the type of motion (for example agile and fast versus precise and slower). These would require not only the physical ability to perform the motor task but also the requisite cognitive ability to adapt based on gameplay scenario (Stuhr et al., 2018). Therefore, interventions can be designed such that improving facets of gameplay performance leads to improved strength, balance, cognition etc., by relating them with modifiable movement signatures (Pacheco et al., 2020) characterized using wearables (Kappen et al., 2019).

The number of participants recruited in the study were sufficient for estimating the movement metrics that were reported in Figure 1. With more than 15 participants, the sample mean of standard deviation for the movement metrics – CoV of AV and ROM, begins to converge, as shown in Figure 2 for participants playing the Fruit Ninja game. Figure 2 shows the distribution of standard deviation of the CoV movement metrics for the Fruit Ninja game (from week 4, left hand sensor). The standard distribution is computed by drawing N participants, where N ranges from 5 to 20 participants. This draw of N participants is repeated 10 times to obtain the distribution shown in Figure 2. While it is challenging to recruit participants to take part in community-centric studies, gamification and social engagement can be good motivators to drive up recruitment for obtaining sufficient participants.

Our study provides a framework for successfully implementing digital health technology in a community setting. Our approach tackled issues on a) trust deficit: by engaging and training health coaches such that they are able to share insights during gameplay, b) resource equitability: by deploying affordable DHT (such as sensors and App) together with grassroots level community engagement allowed sharing of resources thereby ensuring cost-effectiveness, c) technology anxiety: by engaging health coaches to harness on movement behaviour offered by the DHTs to personalize their training regimen for individual participant, d) motivation, and interest: by ensuring flexibility in terms of single vs multiplayer settings during the entire training regimen boosted player confidence, motivation and generated interest that was sustainable during the course of entire training period. In future, studies can directly embed our approach or expand it further to include more than one community centre ensuring competitions on an inter-regional basis.

Further improvements to this study in addressing components related to body function (range of motion, speed of movement for balance etc.), environment (participatory), and personal (age-friendly) factors could support the development of a bio-psycho-social model for healthy ageing (World Health Organization, 2001). In the future, this study would be scaled to a larger participant pool over a longer period with greater community engagement. This will enable further assessment of scalability as well as the longitudinal evaluation of (1) adherence of older adults to physical activity and (2) the efficacy of targeted interventions (exergame design and improvements in health outcomes).

## 5. Conclusions

In this paper, the acceptance and motivation of older adults towards SingaporeWALK, a community-centric, DHT-enhanced solution for supporting healthy ageing is evaluated. The conclusions are as follows:

- Participatory and gamified use of DHTs improved motivation of older adults to be physically active as well as their emotional, social, psychological, and physical health.
- Use of wearables enabled assessment of movement in older adults during exergaming in an unobtrusive manner to provide near real-time feedback.
- Community-centric exergaming significantly improved social and emotional well-being of participants

In the future, interventions can be tailored with specific exergames to target modifiable risk factors for frailty and falls in older adults supported by assessments using wearables.

## Data Availability

Data collected during the study, including de-identified participant data, will be made available on reasonable request, by contacting the corresponding author.

## Conflicts of Interest

All the authors declare no competing interests

## Declaration of generative AI and AI-assisted technologies in the writing process

During the preparation of this work the author(s) used OpenAI-ChatGPT and Grammarly to perform minor edits of the manuscript text. After using this tool/service, the author(s) reviewed and edited the content as needed and take(s) full responsibility for the content of the publication.

## Notes

### Competing Interest Statement

The authors have declared no competing interest.

### Funding Statement

This research was supported by the Intra-CREATE Seed Collaboration Grant (NRF2021-ITS009-0012) and the National Research Foundation, Prime Minister's Office, Singapore under its Campus for Research Excellence and Technological Enterprise (CREATE) programme. The research was conducted at the Future Health Technologies programme and Nanyang Technological University. The Future Health Technologies programme was established collaboratively between ETH Zurich and the National Research Foundation Singapore.

### Author Declarations

Ethical approval for this study was granted by the Nanyang Technological University - Institutional Review Board (IRB-2022-734). Written informed consent was obtained from all participants prior to their involvement in the study. All procedures adhered to the Declaration of Helsinki.

