## Supplementary Materials for "Leveraging community-centric digital health technology to sustain healthier living – inferences from a wearables-based exergaming program"

### Supplementary Material

### A. SingaporeWALK app

SingaporeWALK app is an application developed to promote health and well-being, specifically targeting older adults. The app's design incorporated digital health technologies, such as wearables and exergames. The user interface was crafted to be intuitive and accessible, recognizing that many older adults may not be technologically savvy. The app comprises of following key components:

- Physical Activity Tracker: This feature enables participants to visualize near-real time data related to their arm swing and limb movements, collected from wearables during exergaming and physical activity sessions.
- Nutrition Tracker: Older adults are prompted to monitor their daily dietary habits through the SingaporeWALK app by logging their milk, water, and meal consumption details.
- Well-Being Reporting: Participants regularly report their psychological, social, and emotional well-being through the SingaporeWALK app. The assessment employs the Mental Health Continuum-Short Form (MHC-SF), consisting of three items measuring emotional well-being, six items evaluating psychological well-being, and five items dedicated to social well-being.

While wearables capture movement data in real-time, this data is relayed via a Bluetooth communication to a device such as an iPad using a data collection application for further storage and analysis. With SingaporeWALK, data from wearables is relayed to the iPad, stored locally, and analysed on-the-fly to provide visual feedback to participants related to mobility outcomes such as number of limb movements performed during the game and speed of movement.

SingaporeWALK, in addition to being a data collection app, also serves as an analysis and visualization tool. The app enables the users (participant/health coach) to visualize mobility outcomes calculated by analysing the captured wearables data in a near real-time manner. Information such as number of limb movements performed and force (acceleration) of movement are shown to the participants after a gameplay session. Participants are encouraged to discuss these metrics and their performance with peers that serve as motivation to perform better at the game, socialize and adhere to physical activity.

The app also functions as an interface to collect participant-specific information such as questionnaire data and other agreed-upon personal data. The app is used for an interviewer-administered survey where health coaches collected data on individuals' (a) psychological, (b) social, and (c) emotional well-being each week after the training session. Health coaches also record data about participants' nutrition for the past week using the app. With this information, the health coach would work collaboratively with participants to identify a few lifestyle modifications (in physical activity, well-being, and nutrition) in the coming week.

Therefore, SingaporeWALK is an essential development that serves not only as a data collection platform but also a data visualization and communication tool to support the participants and health coaches.

### B. Exergames

*Fruit Ninja*

Engaging in this game involves moving your arms to slice fruit, effectively exercising the upper limbs and enhancing speed. Fruit Ninja, requires the participants to slice fruits by moving their arms quickly, thereby enhancing speed. The fruits appear on-screen at no set frequency (randomly), at random locations and in random cluster sizes (single or multiple fruits can appear at the same time) with multiple clusters also appearing on screen sometimes. This would require the participant to not only move fast with great range but also to do so with significant repertoire of movements as required to maximize points scored.

*Piano Step*

Playing Piano Steps entails stepping on piano keys, which provides an excellent workout for the lower limbs while also improving balance. In the game, participants have to jump/kick (press down on) on a highlighted piano key with one leg. This game provides a good workout to the lower limbs and improves balance. The keys are spatially pre-determined while the order in which they are highlighted is random. Additionally, the game is split into two bouts. In a slow bout, keys are highlighted at random but slowly across the keyboard. In the second faster bout, the same keys are highlighted repeatedly but at a faster speed, so the participant must jump faster.

*Arctic Punch*

By waving your arms to break ice cubes, Arctic Punch effectively targets the upper limbs and enhances power. This game relies on punching ice blocks that appear on-screen. This game effectively targets the upper limbs and enhances power. The ice blocks in this game mostly appear one-at-a-time but at pre-determined locations (consistent spatially) spanning the entire screen. The time interval between the blocks appearing on screen is also nearly consistent (frequency consistent). The only variation is in the order the blocks appear at the pre-determined locations. Thereby, the game requires quick movement and ability to reach all blocks (range of movement) consistently.

### C. Statistics of movement characteristics over the four weeks of gameplay

Statistics of movement characteristics calculated with data collected using the wearables

| **Game** | **Group** | **Week** | **ROM_mean (°)** | | **AV_mean (°/second)** | | **ROM_cov** | | **AV_cov** | |
| --- | --- | --- | --- | --- | --- | --- | --- | --- | --- | --- |
|  |  |  | **Mean** | **Std Dev** | **Mean** | **Std Dev** | **Mean** | **Std Dev** | **Mean** | **Std Dev** |
| Piano Step | Overall | 1 | 52.80 | 36.81 | 98.39 | 36.61 | 0.36 | 0.12 | 0.48 | 0.20 |
|  |  | 2 | 24.08 | 6.49 | 64.36 | 19.73 | 0.27 | 0.07 | 0.34 | 0.10 |
|  |  | 3 | 24.28 | 8.35 | 66.21 | 24.10 | 0.27 | 0.08 | 0.33 | 0.10 |
|  |  | 4 | 25.03 | 7.47 | 66.71 | 20.94 | 0.29 | 0.07 | 0.34 | 0.11 |
|  | GroupA | 1 | 53.03 | 27.90 | 105.58 | 37.71 | 0.37 | 0.06 | 0.54 | 0.17 |
|  |  | 2 | 25.30 | 6.35 | 64.31 | 18.36 | 0.28 | 0.06 | 0.35 | 0.09 |
|  |  | 3 | 25.00 | 7.76 | 65.56 | 25.27 | 0.30 | 0.07 | 0.37 | 0.08 |
|  |  | 4 | 27.42 | 6.82 | 69.91 | 21.91 | 0.32 | 0.08 | 0.40 | 0.09 |
|  | GroupB | 1 | 52.65 | 41.91 | 93.79 | 35.65 | 0.36 | 0.14 | 0.45 | 0.20 |
|  |  | 2 | 23.17 | 6.54 | 64.39 | 20.99 | 0.26 | 0.07 | 0.33 | 0.10 |
|  |  | 3 | 23.71 | 8.89 | 66.74 | 23.55 | 0.25 | 0.08 | 0.30 | 0.10 |
|  |  | 4 | 23.13 | 7.52 | 64.18 | 20.17 | 0.26 | 0.06 | 0.30 | 0.10 |
| Fruit Ninja | Overall | 1 | 242.28 | 297.08 | 328.48 | 151.98 | 0.47 | 0.20 | 0.46 | 0.18 |
|  |  | 2 | 63.04 | 35.11 | 213.28 | 92.77 | 0.53 | 0.25 | 0.44 | 0.21 |
|  |  | 3 | 59.75 | 38.39 | 197.01 | 92.85 | 0.52 | 0.16 | 0.46 | 0.21 |
|  |  | 4 | 51.28 | 27.47 | 182.79 | 74.09 | 0.56 | 0.18 | 0.47 | 0.18 |
|  | GroupA | 1 | 175.33 | 147.63 | 327.04 | 149.53 | 0.51 | 0.22 | 0.48 | 0.18 |
|  |  | 2 | 64.38 | 33.32 | 217.27 | 84.75 | 0.53 | 0.24 | 0.45 | 0.22 |
|  |  | 3 | 49.83 | 16.79 | 170.75 | 55.37 | 0.57 | 0.12 | 0.53 | 0.20 |
|  |  | 4 | 43.67 | 14.61 | 167.59 | 45.81 | 0.61 | 0.16 | 0.50 | 0.17 |
|  | GroupB | 1 | 289.53 | 362.90 | 329.49 | 155.91 | 0.45 | 0.19 | 0.44 | 0.18 |
|  |  | 2 | 62.01 | 36.95 | 210.20 | 99.81 | 0.53 | 0.26 | 0.43 | 0.22 |
|  |  | 3 | 67.69 | 48.20 | 218.02 | 110.90 | 0.49 | 0.18 | 0.41 | 0.20 |
|  |  | 4 | 57.38 | 33.52 | 194.95 | 89.57 | 0.52 | 0.19 | 0.44 | 0.19 |
| Arctic Punch | Overall | 1 | 132.03 | 77.85 | 203.96 | 66.59 | 0.69 | 0.15 | 0.68 | 0.15 |
|  |  | 2 | 78.15 | 20.74 | 146.35 | 47.38 | 0.57 | 0.11 | 0.66 | 0.12 |
|  |  | 3 | 81.10 | 21.91 | 158.01 | 41.07 | 0.58 | 0.12 | 0.62 | 0.12 |
|  |  | 4 | 75.34 | 21.91 | 149.65 | 39.09 | 0.57 | 0.11 | 0.61 | 0.11 |
|  | GroupA | 1 | 123.44 | 57.97 | 196.42 | 60.22 | 0.74 | 0.13 | 0.69 | 0.13 |
|  |  | 2 | 81.91 | 19.10 | 147.80 | 44.67 | 0.54 | 0.13 | 0.63 | 0.10 |
|  |  | 3 | 86.71 | 20.36 | 161.78 | 34.52 | 0.51 | 0.10 | 0.57 | 0.10 |
|  |  | 4 | 78.77 | 23.67 | 150.99 | 37.93 | 0.53 | 0.09 | 0.59 | 0.09 |
|  | GroupB | 1 | 138.10 | 89.64 | 209.28 | 71.13 | 0.66 | 0.15 | 0.68 | 0.16 |
|  |  | 2 | 75.32 | 21.75 | 145.25 | 50.00 | 0.59 | 0.10 | 0.68 | 0.13 |
|  |  | 3 | 76.61 | 22.40 | 154.99 | 46.01 | 0.63 | 0.11 | 0.66 | 0.11 |
|  |  | 4 | 72.60 | 20.39 | 148.58 | 40.61 | 0.60 | 0.12 | 0.63 | 0.12 |

### D. Significance test results - wearables

**CoV**

GAME - Arctic Punch - For week 1 versus week 2, ROM only, the p_value is: 3.722e-06

GAME - Arctic Punch - For week 1 versus week 2, AV only, the p_value is: 0.255

GAME - Arctic Punch - For week 1, Group A versus Group B, ROM only, the p_value is: 0.014

GAME - Arctic Punch - For week 1, Group A versus Group B, AV only, the p_value is: 0.934

GAME - Arctic Punch - For week 2, Group A versus Group B, ROM only, the p_value is: 0.150

GAME - Arctic Punch - For week 2, Group A versus Group B, AV only, the p_value is: 0.414

GAME - Arctic Punch - For week 3, Group A versus Group B, ROM only, the p_value is: 0.001

GAME - Arctic Punch - For week 3, Group A versus Group B, AV only, the p_value is: 0.012

GAME - Arctic Punch - For week 4, Group A versus Group B, ROM only, the p_value is: 0.080

GAME - Arctic Punch - For week 4, Group A versus Group B, AV only, the p_value is: 0.326

GAME - Fruit Ninja - For week 1 versus week 2, ROM only, the p_value is: 0.038

GAME - Fruit Ninja - For week 1 versus week 2, AV only, the p_value is: 0.234

GAME - Fruit Ninja - For week 1, Group A versus Group B, ROM only, the p_value is: 0.153

GAME - Fruit Ninja - For week 1, Group A versus Group B, AV only, the p_value is: 0.528

GAME - Fruit Ninja - For week 2, Group A versus Group B, ROM only, the p_value is: 0.808

GAME - Fruit Ninja - For week 2, Group A versus Group B, AV only, the p_value is: 0.634

GAME - Fruit Ninja - For week 3, Group A versus Group B, ROM only, the p_value is: 0.064

GAME - Fruit Ninja - For week 3, Group A versus Group B, AV only, the p_value is: 0.003

GAME - Fruit Ninja - For week 4, Group A versus Group B, ROM only, the p_value is: 0.041

GAME - Fruit Ninja - For week 4, Group A versus Group B, AV only, the p_value is: 0.105

GAME - Piano Step - For week 1 versus week 2, ROM only, the p_value is: 8.153e-05

GAME - Piano Step - For week 1 versus week 2, AV only, the p_value is: 1.523e-06

GAME - Piano Step - For week 1, Group A versus Group B, ROM only, the p_value is: 0.254

GAME - Piano Step - For week 1, Group A versus Group B, AV only, the p_value is: 0.101

GAME - Piano Step - For week 2, Group A versus Group B, ROM only, the p_value is: 0.263

GAME - Piano Step - For week 2, Group A versus Group B, AV only, the p_value is: 0.445

GAME - Piano Step - For week 3, Group A versus Group B, ROM only, the p_value is: 0.018

GAME - Piano Step - For week 3, Group A versus Group B, AV only, the p_value is: 0.019

GAME - Piano Step - For week 4, Group A versus Group B, ROM only, the p_value is: 0.029

GAME - Piano Step - For week 4, Group A versus Group B, AV only, the p_value is: 0.001

**Mean**

GAME - Arctic Punch - For week 1 versus week 2, ROM only, the p_value is: 2.941e-07

GAME - Arctic Punch - For week 1 versus week 2, AV only, the p_value is: 7.2641e-07

GAME - Arctic Punch - For week 1, Group A versus Group B, ROM only, the p_value is: 0.8041

GAME - Arctic Punch - For week 1, Group A versus Group B, AV only, the p_value is: 0.3081

GAME - Arctic Punch - For week 2, Group A versus Group B, ROM only, the p_value is: 0.244

GAME - Arctic Punch - For week 2, Group A versus Group B, AV only, the p_value is: 0.949

GAME - Arctic Punch - For week 3, Group A versus Group B, ROM only, the p_value is: 0.036

GAME - Arctic Punch - For week 3, Group A versus Group B, AV only, the p_value is: 0.222

GAME - Arctic Punch - For week 4, Group A versus Group B, ROM only, the p_value is: 0.345

GAME - Arctic Punch - For week 4, Group A versus Group B, AV only, the p_value is: 0.446

GAME - Fruit Ninja - For week 1 versus week 2, ROM only, the p_value is: 2.245e-08

GAME - Fruit Ninja - For week 1 versus week 2, AV only, the p_value is: 1.620e-07

GAME - Fruit Ninja - For week 1, Group A versus Group B, ROM only, the p_value is: 0.515

GAME - Fruit Ninja - For week 1, Group A versus Group B, AV only, the p_value is: 0.961

GAME - Fruit Ninja - For week 2, Group A versus Group B, ROM only, the p_value is: 0.900

GAME - Fruit Ninja - For week 2, Group A versus Group B, AV only, the p_value is: 0.634

GAME - Fruit Ninja - For week 3, Group A versus Group B, ROM only, the p_value is: 0.580

GAME - Fruit Ninja - For week 3, Group A versus Group B, AV only, the p_value is: 0.197

GAME - Fruit Ninja - For week 4, Group A versus Group B, ROM only, the p_value is: 0.528

GAME - Fruit Ninja - For week 4, Group A versus Group B, AV only, the p_value is: 0.467

GAME - Piano Step - For week 1 versus week 2, ROM only, the p_value is: 2.606e-09

GAME - Piano Step - For week 1 versus week 2, AV only, the p_value is: 5.861e-08

GAME - Piano Step - For week 1, Group A versus Group B, ROM only, the p_value is: 0.291

GAME - Piano Step - For week 1, Group A versus Group B, AV only, the p_value is: 0.397

GAME - Piano Step - For week 2, Group A versus Group B, ROM only, the p_value is: 0.198

GAME - Piano Step - For week 2, Group A versus Group B, AV only, the p_value is: 0.794

GAME - Piano Step - For week 3, Group A versus Group B, ROM only, the p_value is: 0.272

GAME - Piano Step - For week 3, Group A versus Group B, AV only, the p_value is: 1.0

GAME - Piano Step - For week 4, Group A versus Group B, ROM only, the p_value is: 0.071

GAME - Piano Step - For week 4, Group A versus Group B, AV only, the p_value is: 0.714

### E. Significance test results – Questionnaires

| (a) All participants | | | | |
| --- | --- | --- | --- | --- |
|  | **feature** | **mean_change** | **se_change** | **p_value** |
| **0** | phy_health | 0.308 | 0.250 | 0.083 |
| **1** | emo_health | 1.256 | 0.266 | 0.000 |
| **2** | social_health | 0.431 | 0.251 | 0.021 |
| **3** | psy_health | 0.378 | 0.291 | 0.080 |
| **4** | Int_exc | 0.872 | 0.246 | 0.000 |
| **5** | Int_game | 0.244 | 0.227 | 0.142 |
| **6** | Int_wearable | 0.269 | 0.236 | 0.103 |
| **7** | Int_app | 1.013 | 0.188 | 0.000 |
| **8** | PU | 0.606 | 0.275 | 0.003 |
| (b) Group A (qualified for the Fun Day) | | | | |
|  | **feature** | **mean_change** | **se_change** | **p_value** |
| **0** | phy_health | 0.583 | 0.249 | 0.053 |
| **1** | emo_health | 1.194 | 0.268 | 0.005 |
| **2** | social_health | 0.133 | 0.222 | 0.502 |
| **3** | psy_health | 0.292 | 0.243 | 0.229 |
| **4** | Int_exc | 0.833 | 0.276 | 0.021 |
| **5** | Int_game | 0.194 | 0.178 | 0.263 |
| **6** | Int_wearable | 0.389 | 0.296 | 0.199 |
| **7** | Int_app | 1.056 | 0.211 | 0.007 |
| **8** | PU | 0.479 | 0.253 | 0.092 |
| (c) Group B (did not qualify for the Fun Day) | | | | |
|  | **feature** | **mean_change** | **se_change** | **p_value** |
| **0** | phy_health | 0.071 | 0.231 | 0.739 |
| **1** | emo_health | 1.310 | 0.263 | 0.000 |
| **2** | social_health | 0.686 | 0.251 | 0.019 |
| **3** | psy_health | 0.452 | 0.325 | 0.182 |
| **4** | Int_exc | 0.905 | 0.216 | 0.005 |
| **5** | Int_game | 0.286 | 0.262 | 0.291 |
| **6** | Int_wearable | 0.167 | 0.163 | 0.282 |
| **7** | Int_app | 0.976 | 0.165 | 0.001 |
| **8** | PU | 0.714 | 0.288 | 0.016 |
